# Integrating primary care and social services for older adults with multimorbidity: A qualitative study

**DOI:** 10.1101/2021.01.30.21250563

**Authors:** Hajira Dambha-Miller, Glenn Simpson, Lucy Hobson, Doyinsola Olaniyan, Sam Hodgson, Paul Roderick, Simon DS Fraser, Paul Little, Hazel Everitt, Miriam Santer

**Affiliations:** School of Primary Care, Population Sciences and Medical Education (PPM), University of Southampton; General Medicine Department. Hinchingbrooke Hospital, North West Anglia NHS Trust

## Abstract

**Background:** Growing demand from an ageing population, chronic preventable disease and multimorbidity has resulted in complex health and social care needs requiring more integrated services. Integrating primary care with social services could more efficiently utilise resources, and improve experiences for patients, their families and carers. There is limited evidence on progress including key barriers and drivers of integration to inform large-scale national change.

**Aim:** To elicit stakeholder views on drivers and barriers of integrated primary care and social services. and highlight opportunities for successful implementation.

**Design and setting:** A qualitative interview study.

**Method:** Semi-structured interviews with maximum variation sampling to capture stakeholder views across services and professions.

**Results:** Thirty-seven interviews were conducted across England including GPs, nurses, social care staff, commissioners, local government, voluntary and private sectors, patients and carers. Drivers of integration included groups of like-minded individuals supported by good leadership, expanded interface roles to bridge gaps between systems and co-location of services. Barriers included structural and interdisciplinary tension between professions, organisational self-interest and challenges in record-sharing.

**Conclusions:** Drivers and barriers to integration identified in other contexts are also present in primary care and social services. Benefits of integration are unlikely to be realised if these are not addressed in the design and execution of new initiatives. Efforts should go beyond local and professional level change to include wider systems and policy-level initiatives. This will support a more systems-wide approach to integrated care reform, which is necessary to meet the complex and growing needs of an ageing multimorbid population.

## Introduction

The ageing population is increasing and by 2035 the absolute number of people aged 65 years or older in England is projected to rise by over 48%.[1] Living with multiple health conditions is more common with increasing age. At least 54% of the UK population over 65 years is currently living with two or more long term health conditions (multimorbidity), and this will exceed 66% over the next decade.[2] Multimorbidity is associated with reduced functional status, increased healthcare utilisation, longer hospital stays and more complex psychosocial needs. The implication is substantial health and social support for personal care needs, assistance with mobility, housing or financial support alongside disease management. Consequently, demand for health and social services is likely to increase further, adding to the strain on existing services. To address this pressing policy issue, the NHS Long Term Plan and the UK Government’s ongoing social care policy review proposed more integrated health and social services.[3,4] As an estimated 45% of the primary care workload includes managing social care needs, integration with social services could potentially release capacity in primary care, reduce duplication of work, increase efficiency and improve experiences for patients and their families.[5] Integration can contribute to higher patient satisfaction, better physical and mental health outcomes, and a recent review of 27 integrated care programmes for adults with chronic diseases, reported increased treatment adherence and lower health costs.[6] An umbrella review of 50 systematic reviews suggested that integrating health and social care can limit costs by facilitating reduced emergency admissions, readmissions, and encourage care within patient’s homes for as long as possible.[7]

The structure and funding of social care, however, markedly differs from primary and secondary care. In England, 152 local authorities manage social care, with statutory responsibilities for assessing individuals’ needs, commissioning services and safeguarding. Over 90% of adult social services are provided by private and voluntary organisations.[8] Unlike healthcare, social care is not a universally free service, rather it is funded through a mixture of central government grants, council tax, business rates and charging people who can afford to pay. These structural and funding differences between the health and social care sectors, present a complex challenge in the effort to integrate services. Furthermore, there is limited conceptual understanding of what closer alignment between these services might look like in practice and integrated care models have been implemented with limited evidence. For example, ‘Vanguard’ sites were established in 2015 to test models of integrated care, followed by interventions such as ‘social prescribing’, ‘care navigation’ and the more recent Integrated Care Systems programme; none of which underwent thorough evaluation prior to national roll-out.[9,10] To date, there remains limited evidence, especially from qualitative studies, investigating progress towards integration specific to primary care and social services in England. Our study aims to address this gap in the evidence base. We explored the topic from the perspective of stakeholders to elicit respondents’ views on progress, drivers of and barriers to existing integration strategies. We highlight examples of successful implementation and provide insights that could inform efforts to achieve closer integration.

## Method

### Design

While we approach this research from a primary care and social care perspective, it is evident that any study of integration must be framed within a systems-wide context [14], which takes account of all dimensions of health and social care. Using this holistic approach, we conducted a qualitative semi-structured interview study with key stakeholders delivering and using these services.

### Recruitment and sampling

We employed purposive sampling to capture a range of participant views. Sixty-three individuals were sent an email invitation; thirty-seven responded and were interviewed. We recruited participants across England from key sectors; primary care, adult social services, secondary care, third sector providers, care home sector, public health, housing, health and wellbeing board, patients and carers. Given the complex structure of health and social care, an iterative and proactive recruitment approach was necessary. We used a snowball sampling technique from the initial round of interviews to identify further participants.

### Data collection

Telephone interviews were conducted between June-September 2020, each lasting between 30 and 60 minutes. An interview schedule was designed (figure 1) covering broad questions to enable similar subjects to be addressed across the sample. A flexible approach ensured related subjects of importance could be raised. Design of the interview schedule was informed by the study aim, an earlier scoping review, expertise of team members and then tested prior to use. Later interviews did not identify significant additional codes, views or experiences so we concluded that data saturation was achieved. Interviews were audio-recorded, transcribed verbatim and anonymised.

### Data analysis

We employed an iterative form of inductive thematic analysis. The first stage involved becoming familiar with the data.[11]. Three members of the team (HDM, GS and SH) independently coded a sample of transcripts from the first round of interviews, then met to discuss initial interpretations until consensus was reached leading to the formulation of a coding framework. Subsequent rounds of coding were conducted with further iterative refinement of the framework. We identified recurring patterns in the data leading to the development of themes. From this, an interpretative narrative of the data was written. Throughout the analytical process, a form of constant comparative analysis was used to identify key differences or similarities in the data, between professions, sectors or geographies, etc. We also searched for alternative perspectives that may have challenged our interpretations. This process of deviant case analysis reduced the risk of bias and added rigour to our analytical conclusions.

QSR NVivo software (version 12) was used to manage the data and the Consolidated Criteria for Reporting Qualitative Research (COREQ) checklist guided reporting.

### Data availability

Data are available from authors with a reasonable request.

## Results

We interviewed 37 people; 23 females and 14 males. Participants comprised seven patients/carers and 30 professionals, from across care sectors and regions of England. (Table 1) Our analysis identified three overarching themes and additional subthemes which are discussed narratively below with supporting quotes.

### Theme 1: Facilitators of primary care and social services integration

Participants highlighted factors facilitating integrated care for older people experiencing multimorbidity as follows:

#### Individuals and teams driving integration

Participants identified the role that key individuals or teams of like-minded collaborators can play as innovators and drivers of integration; *‘there is a brilliant geriatrician …who had this proactive approach and worked very well also with her colleagues in GP practices …She was trying to coordinate things across the system, and …it really works well. A lot of it though is dependent on charismatic individuals*.*’ (Participant 14, Public Health)*

Interviewees credited individuals driving integration with recognising the benefits of empowering others and creating a culture that encourages initiative among frontline professionals, enabling them to develop joined-up solutions; *‘it’s just the people on the ground feeling they’ve got the trust, and the freedom and the expectation to come up with ideas when they’re seeing that things could work better* …*that really comes from Dr K empowering me and my team, and those around her’ (Participant 18, Primary Care/Community Services)*

Team building was identified as essential to integration, described by an interviewee as an incremental developmental process, building iteratively; *‘We built things at a steady pace. …it’s constant work …started with a small core district nurse GP Social Prescriber and our Hub Coordinator Nurse, and we’ve built from there. So rather than waiting for the whole set to be ready, we’ve got started, we’ve built a good, strong core team. Then, social care were willing to come into that functional group. …mental health have come in*.*’ (Participant 19, Primary Care)*

Some participants stressed that integration requires leadership across all scales and sectors of health and social care, especially to ensure resources align with demand; *‘having the willingness of the right people at the right level to say: Okay, so maybe the capacity is in the wrong place*.*’ (Participant 24, Secondary Care)*

### Interface roles

Participants identified the importance of non-clinical and clinical co-ordination roles, with various titles such as Care Navigators or Integrated Care Co-ordinators who work at the intersections where primary care, secondary care and social services meet; *‘in the GP surgery, they had their own team who were involved more with social issues …and they called them health coordinators. It was one of the workers there helped me [an Adult Social Worker] to organise sorting out his [an older adult client] house, because it was in a bit of a state*.*’ (Participant 8, Adult Social Care)*

Operationally, these interface roles were viewed as critical in facilitating integration among service providers by bridging gaps across sectoral boundaries; *‘social prescribers are the lynchpin of linking primary [care] with adult services …the enabler that gives that bit of confidence …the bridge between the two*.*’ (Participant 28, Voluntary Sector)*

Care co-ordinators were described as crucial in addressing the everyday social care and psycho-social needs of older people experiencing multimorbidity, once discharged into community settings; *‘someone had gone home, a daughter had gone on holiday to Italy with the keys* …*The care navigator said: With the say-so of the patient …can we get a new lock put on your door with a new set of keys, and we can discharge you home? …actually, non-clinically, looking at the issue of saying: Okay, you’ve sorted out the clinical, let me sort out the social and community aspect*.*’ (Participant 33, Voluntary/Statutory Sector)*

Care co-ordinators were perceived as system navigators by carers and patients, providing support and advice when navigating the complex systems of health and social care; *‘There’s a real need for maybe an elderly care coordinator* …*Not necessarily with every GP practice, but within a hub of GP practices that you have somebody that’s responsible for the elderly people in your community. …and maybe trying to ensure that they are in touch with the people that they need to be’. (Participant 3, Carer)*. Another carer said; *‘when you’ve got four or five different things going on, you think …if there was just one person and we spoke to them and said: can this happen? That would make a massive difference*.*’* (*Participant 2, Carer)*

Having in place a layer of professionals located at potential ‘pinch points’ in the systems of health and social care was identified as not only significant in terms of reducing delays and blockages, but also in enabling seamless care transition across sectoral boundaries; *‘some GPs are incredibly helpful, some aren’t. Some won’t share any information with us [Adult social care]. Every surgery has got a clinical coordinator. If you’ve got a slightly risky discharge [from an acute hospital], we would phone them and say: Mrs Smith is coming home with a four times daily care package, can you just pop out and see them? …If they’ve got any concerns about their clients who are admitted into hospital, they’re very quick to phone us and say, give us the back story*.*’ (Participant 11, Adult Social Care)*

### Co-location and collaboration

Participants identified co-location as a spatial and social enabler of integration. This concept is understood as a shared working environment where professionals from various disciplines can interact and collaborate; *‘[To improve co-ordination of hospital discharge among partners]* …*I [Integrated Discharge Service Lead] tried to get their input into how we could change things in their environments, as well as processes. Then I changed it, and we then just had a monthly …we all stopped for an hour and we did all sorts of things. That was mainly just to try and get them to mix, talk to each other from different organisations. …That was helpful, just so that they could then appreciate where each other were …and also for me, it then helps to see how each one works differently*.*’ (Participant 24, Secondary Care)*

Shared working spaces were identified as facilitating inter-professional relationship building and bonds of trust, which are essential to establishing sustainable, integrated working arrangements; *‘it is about those, literally, working in the same offices. I think it’s also about relationships …if somebody lands on somebody’s door, we’re now saying: actually, it might not be the right place for us, but actually we know who can support you and where you can go*.*’ (Participant 18, Primary Care/Community Services)*

The emergence of a shared multi-disciplinary team ethos in co-located spaces appeared to create an environment that enables professionals to challenge one another and engage in difficult conversations about appropriate options for patient care; ‘*It took probably six to 12 months, I would say, for us to … [become a joined-up inter-professional team]. What we do now, we go into meetings and we really challenge one another, but we do that from the point of view: … I’m not angry with you …I’m just doing my role …It was really difficult at first* …*Now, I think there’s a level of respect there for each other*.*’ (Participant 11, Adult Social Care)*

Translating a vision of integrated working into practice, requires stakeholders to agree a plan of action setting out a modus operandi of how they will collaborate and align activities; *‘I went to visit [a] hospital down in Somerset* … *What they did actually … is they went and sat everybody in a basement for a week from everywhere, all of the organisations, and said: We are not leaving until we’ve come up with a plan to work together. from what I could tell … it has had a huge impact on them as a county*.*’ (Participant 24, Secondary Care)*

### Theme 2: Where integration occurs

Participants highlighted the multi-level nature of health and social care integration, which occurs at and across different scales. Our data suggest that efforts to drive forward integration are mainly focused on two scales. First, there are micro-scale clinical initiatives that aim to join-up care at the point of delivery to the patient; ‘*the acute Trust were really keen on having social work presence at the front door…they ring us [Adult social care] and we’ll be down there within …four hours, is the agreement’. (Participant 11, Adult Social Care)*. Another participant describes; *‘[the GP practice] employs a discharge liaison person to work at …our local hospital …We’ve got that really nice link of somebody …who’s working in the hospital*.*’ (Participant 18, Primary Care/Community Services)*

Second, integration also takes place at the meso-level, often in the form of inter-professional collaboration and joint arrangements between organisations; *‘We all [Adult Social Care] work quite well with mental health because there’s jointly funded posts it’s not just looking at things from one angle, it’s looking at it from, I guess, a more holistic point of view. What it means for the person in their life, rather than what it means for the person with their social care and what it means to them with their medical needs. It’s smoother*.*’ (Participant 12, Adult Social Care)*

*‘I [Senior GP] no longer have just GPs in the practice, but I have paramedics, pharmacists and nurse practitioners, practice nurses. …we’ve got a physio within the practice now -a social prescriber. I think these are major steps forward*.*’ (Participant 21, Primary Care)*

Integration requires mechanisms which operate across scales by bringing together different health and social care sectors. Operationally, integration is delivered through collaborative structures such as multi-disciplinary teams; *‘so, it’s a really good two-way thing. That unplanned admissions team is absolutely essential to the way we [social prescribing community development service] work, and our working together is really crucial. The MDT [multi-disciplinary team] meetings that came alongside that … which was us, the unplanned admissions team, district nurses, our discharge liaison*.*’ (Participant 18, Primary Care/Community)*

### Theme 3: Tensions

A number of tensions in progress towards greater integration were identified. Structural tensions were an inherent feature of the complex multi-layered configuration of health and social care; ‘*On the one hand, the system is not really well designed to support that integrated working. So, someone in the hospital … They’ll really concentrate as hard as they can on that period but then once that’s finished, they move on to the next person. Even the language and even the funding structures that support that approach, to a lesser or greater degree depending on where you are*.*’ (Participant 30, Voluntary Sector)*

Health and social care are delivered through a series of separate systems and organisations, which in itself is an inhibitor of integration. *‘It’s about problem-solving rather than just retrenching to your own bit and lobbing stones. I think culturally, that’s been quite difficult because our systems are set up quite adversarially in a way, you know, everybody’s got their …own little silo to protect*.*’ (Participant 17, Local Government)*. These separate structures can lead to tensions emerging among organisations. Most frequently, participants identified the tendency of organisations adopting a silo mentality, which emphasises internal priorities over potential benefits arising from external collaboration; *‘there is a huge amount of siloed thinking. The hospitals are very good at protecting their areas of expertise by using NICE guidance. This was about the hospital making sure they kept control of a particular speciality*… *(Participant 21, Primary Care)*

Organisational self-interest and protectionism, which is an institutional response involving organisations protecting their interests and retaining control over specialisms, was identified as a further barrier to integration; *‘I [GP] deal with [a neighbouring hospital trust] quite a bit* … *The systems there are much slicker because you don’t have this territorialism*.*’ (Participant 26, Secondary Care)* Participants also identified poor communication as inhibiting integration, both between and within organisations; *‘there needs to be better communication as well, between the GP surgery and between ourselves (Adult Social Care)… when we have safeguarding concerns and there’s a … professionals meeting -sometimes they don’t turn up*.*… and there’s constant arguments between us and the GP practice, and then it just becomes really draining*.*’ (Participant 9, Adult Social Care)*

Another well-documented tension raised by participants is the inability to share records (e.g. health records/history, prescribing information and personal details) between service providers due to factors such as systems incompatibility and uncertainty over legal requirements relating to information sharing; *‘we have no integration between these different systems. I think this is everybody’s biggest bugbear. So much time would be saved by being able to dive into each other’s medical records and look at what’s going on’ (Participant 26, Secondary Care)*.

For patients, carers and families navigating the series of systems which constitute health and social care provision can be a frustrating challenge; *‘we felt that we were having to speak to so many different people. You’d go to one person and they’d deal with that bit, and the next person would deal with another bit, and another person’ (Participant 2, Carer)*

Participants highlighted how tensions among health and social care actors were playing out across spatial scales; *‘it was about the top down structure. So, the other doublespeak is that they want policies …to be developed from the bottom up, but universally it’s always top down because that’s where the funding decisions come from, and until we truly give the money to (local primary care) networks, for them to absolutely decide what their priorities are, it’s never really going to change*.*’ (Participant 21, Primary Care)*

Some participants argued for more practice- and solutions-based approaches that are localised and emerge at the clinical and professional scales from empowered individuals with the autonomy to act; *‘surely somebody in the top tables are trying to figure out how this can happen; …sometimes it’s just the people on the ground feeling they’ve got the trust and the freedom and the expectation to come up with ideas when they’re seeing that things could work better; rather than thinking, we have to wait for someone on the top table to figure out the answer to something*.*’ (Participant 18, Primary Care/Community Services)*. Another participant said; *‘GP-based care is not the best way forward and actually delegation and making sure that lots of other people do these things, should work well. We just have to ensure that the teams communicate well and that the teams have a feeling of autonomy*.

*My worry is that this [the integration agenda] has been approached in a rather piecemeal fashion*.*’ (Participant 21, Primary Care)*

## Discussion

### Summary of key findings

This study explored stakeholder views on progress, drivers of and barriers to existing integrated care initiatives within primary care and social services in England. Like-minded individuals were often key drivers of integration supported by strong leadership. Interface roles were emphasised to bridge gaps between providers, facilitate seamless service provision and support patients/carers to navigate the complex health and social care infrastructure. Co-location were ‘creative’ spaces enabling formal and informal integration. In practice, integration was mainly focused on micro-level frontline clinical initiatives to facilitate interdisciplinary working among professionals, while concomitantly there were tensions in progressing towards greater systems-wide integration. These tensions were viewed as an inherent feature of health and social care delivery, which was a series of disparate organisational structures and systems where there was limited learning and progress from previously tested models and initiatives.

### Comparison to existing literature

Our results are consistent with earlier work on integration, except few previous studies have examined primary care and social services.[12,13] It is concerning that many of the factors identified as important in this study have been highlighted in previous literature in other areas of integrated care, yet remain poorly applied in newer models of integration. This may well be contributing to the inadequate pace and progress of integrated services. The pivotal role of leaders and leadership in driving and sustaining integrated working are highlighted in earlier studies but are still not sufficiently prioritised in current care models or policy-making.[14,15] Building on the evidence base, we highlight the importance of innovative individuals and teams who, if empowered, can drive forward integration. Our finding that co-location is effective in bringing together professionals from multi-disciplinary backgrounds to deliver integrated care, needs to be considered in light of the recent move towards remote collaboration and virtual meetings due to COVID-19. Going forward, pandemic induced change potentially may accelerate integration in other aspects of electronic/IT-based working such as improved information and records sharing.[16] The role of interface staff such as care-navigators and social prescribers was emphasised by respondents. Our results suggest that for patients/service users and carers, these roles provide a single point of contact that aided navigation of complex care systems and facilitated access to services. This in turn can contribute to a more seamless care transition between services.[9,17] The barriers to integration relate to tensions that have been widely reported, such as the substantial challenges of sharing records across organisational boundaries, poor communication among service providers especially concerning older patients with multimorbidity transitioning between services, and the protectionism evident among organisations and professionals focused on their own specialist interests. [18]

### Strengths and Limitations

A strength of this study is the large sample of participants from a diverse range of professions, regions, sectors and scales of integration including strategic-level management, frontline clinical and non-clinical service providers, alongside service users and their carers. Use of semi-structured interviews enabled open-ended probing and in-depth exploration of participants’ views, allowing us to understand integration more holistically. However, the use of snowball sampling and the study’s reliance on professionals and layperson participants voluntarily opting-in to the research may have introduced an element of self-selection. COVID-19 limited methodological options and given the restrictions imposed on social interaction we had to conduct telephone interviews. This may have been a factor in our sample of patients. It is plausible that different accounts may have been obtained with in-person interviews. Understandably, the responses of participants may have, to some degree, been framed by the impact of the ongoing pandemic on contemporary health and social care practice. It is possible that if this study was conducted before or after the pandemic, our findings may have differed.

### Implications for practice

Our study highlights well-established drivers and barriers to integration that is also present in the primary care and social services context. Progress towards integration is likely to continue at a slow pace if these are not adequately considered in the design and execution of new initiatives. Failure to learn from previous models is concerning. We found that it is essential to harness the potential of dynamic key individuals and/or teams to drive forward integration and our findings add weight to the evidence base on the value of new interface roles. These models could potentially be expanded further, although with the important caveat that any wider rollout has greater efficacy if part of other system-wide processes of integration. Our research reports that efforts to progress integrated working have mainly concentrated on the clinical and professional aspects of integration located at the micro and meso levels of health and social care structures, indicating that integration has, to date, primarily been a bottom-up process. This suggests that there is potential scope to examine whether macro-scale integration can be increased at the higher organisational and strategic level across health and social care, and in doing so, contribute to a more holistic systems-wide approach to reform across England.

## Data Availability

Data used during the current study are available from the corresponding author on reasonable request.

## Acknowledgements

The authors wish to thank all participants and our PPI representatives for their contributions.

## Ethical Approval

All participants provided written informed consent, and ethical approval was obtained from University of Southampton Ethics committee (reference number: 56311).

## Author Contribution

HDM designed the study, wrote the analysis plan, conducted the analysis, drafted and revised the paper. GS contributed to the analysis, drafted and revised the paper. SH, LH and DO contributed to the analysis and revised the paper. MS, HE, SF and PR contributed to the study design and revised the paper. HDM is guarantor.

## Declaration

The lead author affirms that the manuscript is an honest, accurate, and transparent account of the study being reported; that no important aspects of the study have been omitted. The opinions, results, and conclusions reported in this article are those of the authors and are independent of the funding sources.

## References

1 Barnett K, Mercer SW, Norbury M, et al. Epidemiology of multimorbidity and implications for health care, research, and medical education: A cross-sectional study. The Lancet 2012;380:37–43. DOI:10.1016/S0140-6736(12)60240-2

2 Cassell A, Edwards D, Harshfield A, et al. The epidemiology of multimorbidity in primary care: A retrospective cohort study. British Journal of General Practice 2018;68:e245–51. DOI:10.3399/bjgp18X695465

3 The King’s Fund. The NHS long-term plan explained. 2019. https://www.kingsfund.org.uk/publications/nhs-long-term-plan-explained?utm_source=TheKing%27sFundnewsletters%28mainaccount%29&utm_medium=email&utm_campaign=10222788_MKPUB_NHSlong-termplanlongread24012019&utm_content=btnbel (accessed 24 Jan 2019).

4 Jarrett T. Adult social care: the Government’s ongoing policy review and anticipated Green Paper (England). /research-briefings/cbp-8002/ (accessed 23 Nov 2020).

5 Saxe Zerden L de, Lombardi BM, Jones A. Social workers in integrated health care: Improving care throughout the life course. Social Work in Health Care 2019;58:142–9. DOI:10.1080/00981389.2019.1553934

6 Martínez-González NA, Berchtold P, Ullman K, et al. Integrated care programmes for adults with chronic conditions: A meta-review. International Journal for Quality in Health Care 2014;26:561–70. DOI:10.1093/intqhc/mzu071

7 Damery S, Flanagan S, Combes G. Does integrated care reduce hospital activity for patients with chronic diseases? An umbrella review of systematic reviews. BMJ Open. 2016;6. DOI:10.1136/bmjopen-2016-011952

8 Humphries R. Integrated health and social care in England - Progress and prospects. Health Policy 2015;119. DOI:10.1016/j.healthpol.2015.04.010

9 Husk K, Elston J, Gradinger F, et al. Social prescribing: Where is the evidence? British Journal of General Practice 2019;69:6–7. DOI:10.3399/bjgp19X700325

10 Health and social care integration – National Audit Office. http://www.nao.org.uk/report/health-and-social-care-integration/ (accessed 15 Feb 2017).

11 Braun V, Clarke V. Using thematic analysis in psychology. Qualitative Research in Psychology 2008;3:77–101. DOI:10.1017/CBO9781107415324.004

12 Hughes G. Experiences of integrated care: reflections on tensions of size, scale and perspective between ethnography and evaluation. Anthropology and Medicine 2019;26:33–47. DOI:10.1080/13648470.2018.1507105

13 Ling T, Brereton L, Conklin A, et al. Barriers and facilitators to integrating care: Experiences from the English integrated care pilots. International Journal of Integrated Care 2012;12. DOI:10.5334/ijic.982

14 Wistow G. Hope over experience: still trying to bridge the divide in health and social care. Published Online First: 1 August 2017. https://www.ippr.org/ (accessed 24 Nov 2020).

15 Evidence review - integrated health and social care - Institute of Public Care - Oxford Brookes University. https://ipc.brookes.ac.uk/publications/publication_809.html (accessed 24 Nov 2020).

16 Brown L, Tucker C, Domokos T. Evaluating the impact of integrated health and social care teams on older people living in the community. Health Social Care Community 2003;11:85–94. DOI:10.1046/j.1365-2524.2003.00409.x

17 Drinkwater C, Wildman J, Moffatt S. Social prescribing. BMJ (Online) 2019;364. DOI:10.1136/bmj.l1285

18 Maruthappu M, Hasan A, Zeltner T. Enablers and barriers in implementing integrated care. Health Syst Reform. 2015;1:250–256. DOI: 10.1080/23288604.2015.1077301

