## Supplemental table for "Integrating primary care and social services for older adults with multimorbidity: A qualitative study"

**Box 1: Interview schedule**

**A: Interviews with health and social care staff**

1.Describe your role in delivering integrated or joined-up health and social care and the links you have with other professionals and organisations

2. What aspects of integrated or joined-up health and social care have worked well, and why?

3. What aspects of integrated or joined-up health and social care worked less well, and why?

4. What changes would you like to see in these services, in terms of making them more integrated or joined-up?

**B: Interviews with patients, relatives and carers**

1. Please tell me about the reasons why you began using health and social care services?
2. What aspects of these services worked well together for you?
3. What services did not work well?
4. What changes would you like to see in these services, especially in terms of making these services more integrated or joined-up?
