## Supplemental table for "Integrating primary care and social services for older adults with multimorbidity: A qualitative study"

| **ID** | **Sector** | **Region** |
| --- | --- | --- |
| 1. | Carer/relative | Berkshire |
| 2. | Carer/relative | Berkshire |
| 3. | Carer/relative | Oxfordshire |
| 4. | Carer/relative | Northumberland |
| 5. | Relative | Leicester |
| 6. | Relative | London |
| 7. | Patient | Oxfordshire |
| 8. | Local government, Adult Social Care | Wiltshire |
| 9. | Local government, Adult Social Care | Peterborough |
| 10. | Local government, Adult Social Care | Cambridge |
| 11. | Local government, Adult Social Care | Wiltshire |
| 12. | Local government, Adult Social Care | Cambridgeshire |
| 13. | Local government, Department of Community Services | Wiltshire |
| 14. | Local government, Public Health | Hampshire |
| 15. | Local government, Housing Department | Oxfordshire |
| 16. | Local government, Housing Department | Oxfordshire |
| 17. | Local government, Health and Wellbeing Board | Northumberland |
| 18. | Primary Care/Community Services | Somerset |
| 19. | Primary Care | Somerset |
| 20. | Primary Care/Care Navigation | Hampshire |
| 21. | Primary Care | Dorset |
| 22. | Primary Care/Ambulatory care | Oxfordshire |
| 23. | Secondary Care, Community Nursing | Northumberland |
| 24. | Secondary Care, Discharge Service | Somerset |
| 25. | Secondary Care, Care Co-ordination | Northumberland |
| 26. | Secondary Care, Urgent Care Services | Oxfordshire |
| 27. | Voluntary Sector | Leicester |
| 28. | Voluntary Sector | Hampshire |
| 29. | Voluntary Sector | London |
| 30. | Voluntary Sector | London |
| 31. | Voluntary Sector | Northumberland |
| 32. | Voluntary/Statutory Sector | Hampshire |
| 33. | Voluntary/Statutory Sector | Hampshire |
| 34. | Voluntary/Statutory Sector | Hampshire |
| 35. | Care Home Sector | Lancashire |
| 36. | Care Home Sector | Northumberland |
| 37. | Care Home Sector | Yorkshire |
